# Interventions that could mitigate the adverse effects of household overcrowding: A rapid realist review with stakeholder participation from urban contexts in England

**DOI:** 10.1101/2024.09.10.24313301

**Authors:** Kristoffer Halvorsrud, Elizabeth Eveleigh, Mathilda O’Donoghue, Pratima Singh, Rose-Marie McDonald, Marcella Ucci, Jessica Sheringham

**Affiliations:** Research Department of Primary Care and Population Health, University College London (UCL), 1-19 Torrington Place, London WC1E 7HB, UK; Wolfson Institute of Population Health, Queen Mary University of London (QMUL), Yvonne Carter Building, 58 Turner St, London E1 2AB, UK; PPIE member; UCL Institute for Environmental Design and Engineering, Bartlett Faculty of the Built Environment, University College London (UCL), UK

**Keywords:** Housing, Overcrowding, Rapid realist review

## Abstract

Household overcrowding has increased in England. However, there is limited synthesis of evidence about what can be done to reduce the impact of overcrowding on health/well-being.

We undertook a rapid realist review of English language peer-reviewed and grey literature of interventions from comparable settings to urban contexts in England that addressed household overcrowding/health outcomes. A search was conducted (01.06.23) in MEDLINE, EMBASE, Web of Science, SCOPUS.

Two expert panels informed the review. The first comprised individuals with lived experience of overcrowding in London; the second local and regional government representatives from London, Salford and Doncaster (England). Both panels contributed at two stages to guide the scope/literature identification and test/refine programme theories. Final full-text screening and quality appraisal were completed by two independent researchers.

Thirty-one peer-reviewed papers and 27 documents from participating local authorities were included. The peer-reviewed literature, emanating from multiple geographical contexts and of variable study designs and quality, contained 15 evaluated interventions across three categories: Rehousing (n=7 interventions); Home improvements, e.g. renovations/retrofitting (n=6); Co-ordination with healthcare and wider services (combined with home improvements) (n=2). A synthesis of the peer-reviewed literature with expert panel comments and grey literature, identified contexts and mechanisms that could facilitate or hinder achievement of positive health outcomes. There was reluctance to be rehoused elsewhere, with residents fearing the loss of social networks in available properties often located far away from their current homes. Home improvements may alleviate the worst impacts of overcrowding, and residents living in unhealthy conditions can benefit from better healthcare co-ordination.

**Significance for public health:** Reducing the prevalence of overcrowding requires national level and long-term policy changes to increase the supply of affordable homes. Therefore, rehousing will not be a feasible solution in the short-term for many residents. Our rapid realist review illustrates how other interventions such as home improvements and improved healthcare co-ordination/access could address well-being when residents in overcrowded accommodation cannot or do not wish to move. This may require overcrowding to be considered as a council-wide issue that may not be tackled within the housing sector alone, but that will necessitate collaboration with other local authority resources and services such as healthcare in recognition of the wider health impacts of overcrowding. Although the focus for this review is on making recommendations for urban contexts in England, we have also included international peer-reviewed literature and believe our conclusions may be transferable to comparable contexts affected by household overcrowding.

## Introduction

The challenge of accommodating rising population numbers in urban areas with limited space is a common one and not a new phenomenon internationally (1). However, measured against comparable countries, England has somewhat larger numbers of residents per household (2). Moreover, statistics for 2019/20 from the English Housing Survey revealed the highest levels of overcrowding in the country since 1995/96, with 8.7 percent of households in the social rented sector and 6.7 percent in the private rented sector being overcrowded (3, 4). Regional variations were also confirmed in figures from the English Housing Survey (2019/20), with as many as 9.2 percent of London households overcrowded in comparison to 2.5 percent for England as a whole (3, 4). While overcrowding during the COVID-19 pandemic reached 15 percent in London (5). Yet this is not just a London phenomenon and there are pockets of overcrowding throughout England, with differences by ethnicity affecting those from a Black, Pakistani or Bangladeshi ethnicity in particular (6).

There are different definitions to characterise a household as ‘overcrowded’ or ‘crowded’. Generally it occurs when the number of residents exceeds the capacity of the dwelling space available, which may be measured variously depending on the geographical context through e.g. number of rooms, floorspace per person (1). In the UK the ‘bedroom standard’ is commonly applied (although also here regional variations exist (7)). This standard assumes that certain household members need to have their own bedroom, while others according to their age and gender, can share (8). There is a recognition that whether a household is ‘crowded’ as such may not invariably relate to the number of people residing in the dwelling, but also demographic characteristics such as age/gender constellations and their relationships (i.e. an otherwise ‘crowded’ household by quantitative measures, may not be so if for example two adults are a couple) (1).

As amply demonstrated in a relatively recent World Health Organization (WHO) review (9), overcrowded often increases the risk of other housing issues such as environmental hazards (e.g. damp and mould, disrepair or clutter) and the risk for respiratory and gastrointestinal infections. Consequently in numbers from England, 40 percent of overcrowded households had reported significant mould in contrast to only 16 percent of non-overcrowded households (5). Combined with other adverse housing circumstances, overcrowding can contribute to poor physical and mental health, but also have wider socioeconomic ramifications such as poorer educational outcomes due for example to a lack of sleep (9).

Although the evidence base for problems associated with household overcrowding are now relatively well established, there is a lack of current evidence on interventions or measures that may reduce overcrowding or the adverse health outcomes associated with overcrowding. The last systematic review that explicitly addressed the effects of housing interventions on health by Thomson and colleagues (10) was published over 10 years ago. Moreover, its broader scopes – housing interventions in general – did not provide more in-depth coverage or isolation of effects that specifically concerned overcrowding *per se* compared to other and potentially non-overcrowding housing issues (due e.g. to poverty). Although a more recent systematic review on buildings and health (11) includes some recommended interventions, it is primarily concerned with and draws from the evidence base on the associations between buildings and health (i.e. not interventions) and again is not restricted to overcrowding.

The present review aimed to fill this research gap. To achieve this aim, we conducted a rapid realist review (RRR) (12) with participation of key stakeholders from urban contexts in England to identify peer-reviewed literature (including international if providing transferable lessons) as well as grey literature of local mechanisms by which interventions to address or mitigate household overcrowding on health/well-being may be effective. A realist approach helped gain a better understanding of which interventions may show promise in which circumstances, than would have been possible if concentrating on effectiveness alone for a complex issue such as housing overcrowding (12).

## Methods

A protocol was pre-registered on PROSPERO (CRD42023396754). We undertook a rapid realist review (RRR) (12), influenced by RAMESES (Realist And MEta-narrative Evidence Syntheses: Evolving Standards) with the list of items to be reported in a realist synthesis (13) and the page numbers where the items have been reported in the present manuscript indicated in Additional file 1. Similar to a full realist review, an RRR provides a framework to collect and synthesise relevant and sufficient evidence on contexts/mechanisms/outcomes (CMO) to theorise how or why a group of interventions or single interventions could achieve their effects. An RRR is a useful tool to respond to time-sensitive and emerging policy issues such as overcrowding where time/resources are more limited than permitted by a full realist review. In particular, this meant a shorter window for iteration or the possibility of adding further documents as one would typically see in a *full* realist review. Yet, stakeholder involvement (see below) was key to ensuring the review remained relevant (12).

Our principal understanding of whether a household is ‘crowded’ concerns the household members’ experience of not having enough space for daily living or to perform activities as they would have wanted to. We theorised that improvements (mechanisms) to such space in the dwelling or their surrounding environments – either quantitatively in actual physical space or qualitatively as experienced amount of available space – may in many contexts (if e.g. the dwelling is not too structurally confined or damaged to make significant improvements) offset the need to move elsewhere (outcomes). Indeed, moving elsewhere might not be practically feasible within dense urban environments with limited space (context/mechanism) (14), or even viewed as a benefit by residents themselves (outcomes). From previous work (15) we also advocate for a more holistic approach joining up not only housing but also other local authority resources and services such as healthcare (mechanism), recognising the wider health impacts of overcrowding (context/outcomes).

### Stakeholder involvement

In an RRR, stakeholder engagement acquires a more explicit part than in a full realist review to streamline the review process. Expert panels of key stakeholders allow for the review to be guided to and oriented around key available literature and for conclusions to be co-developed (12).

As key stakeholders, we convened expert panels of a) individuals with lived experience of household overcrowding according to our definition above (hereafter: ‘residents’) in London (comprising two separate panels in Tower Hamlets and Islington); and b) local and regional government representatives (hereafter: ‘staff’) across London (Greater London Authority, Tower Hamlets, Newham, Camden and Islington) and non-London authorities (Salford and Doncaster). Residents consisted of Tower Hamlets contacts that had participated in a similar project (by MO and PS) and from an Islington charity for ethnic minority women (Jannaty). As both groups consisted mainly of ethnic minority residents, it reflected their overrepresentation in overcrowding statistics and enabled us to engage seldom heard voices (6). For the staff panels, local authorities with an expressed priority for housing problems were approached through our research team’s professional networks. A representative from either public health with a remit for the built environment, wider determinants or from housing, was sought. We believe the involved authorities provided illustrative cases of overcrowding in urban contexts in England. In a 2018 analysis of London authorities and average occupied floorspace per person (2), Tower Hamlets, Newham, Camden and Islington ranked lowest, second lowest, seventh lowest and eighth lowest, respectively (and the 2021 Census also suggested a higher overcrowding percentage in Newham and Tower Hamlets in particular (16)), while Salford and Doncaster are non-London contexts reportedly also challenged by overcrowding (17, 18).

The stakeholder groups took part in two rounds of expert panels each. In the initial panels, we discussed the nature of overcrowding in the local context, as well as initial ideas and experiences of possible interventions, and any prioritised outcomes in their contexts. These discussions guided the review scope and provided any missing terms to our preliminary search strategy. In the second expert panels, categories of interventions identified from the literature, plus questions arising from gaps or uncertainties in the literature, were shared for validation. We asked the panels how identified evidence resonated with the situation in their local contexts, whether the interventions might be relevant or transferable, if there were any mechanisms for making them work which they felt had not been covered and any contextual barriers that might compromise any observed effects/impacts.

The staff panels (first on 19.04.23 (n=11 participants); second on 26.09.23 (n=7)) were facilitated as one-hour structured online discussions enabling national participation including non-London authorities. The residents’ panels in Tower Hamlets (first on 16.05.23 (n=6 participants); second on 18.12.23 (n=5)) and in Islington (first on 24.05.23 (n=12), second on 08.01.24 (n=6)) were facilitated at local venues associated with the respective recruited community groups (see above). These in-person sessions were longer in duration to facilitate relationship building, familiarisation with the research and translation from non-English languages (e.g. Bengali): initially three hours each to accommodate for crucial background detail in conceptual review stages, while the second panels were two hours each specifically focused on findings and the identified intervention categories.

Panel members were not considered research participants (and the sessions should not be conflated with more traditional focus groups). The panel members were considered experts in overcrowding, either from a lived experience or policy perspective. Their input was sought to determine the direction of the research initially, then interpretation or contextualisation of the findings. Therefore ethical approval was not required. This fits within the remit of a rapid realist review to help streamline the process of literature identification and validation (12), but also more broadly within established realist review frameworks to support, refute or contextualise the evidence base in the interest of theory development (13).

However, panel members were still provided with full details including an information sheet on the purpose of the review and panels and they could withdraw at any time without providing any reason. Permission was requested for panels to be audio-recorded to enable researchers to focus on discussion, which in one case was denied, but permission was given to take notes. Recordings and notes of panels were not subject to analysis of individual responses and we do not report on any individualised experiences or sentiments, but only on views for which consensus was indicated by multiple voices that provided rationale for the review’s focus or added to the contextualisation or interpretation of findings.

### Searches/screening process

PROSPERO was initially searched to verify that the proposed review may indeed fill a research gap and that there were no ongoing and/or significantly overlapping reviews on this topic. Due to the complex nature and corresponding solutions to overcrowding, the electronic databases used (MEDLINE, EMBASE, Web of Science, SCOPUS) accounted for multiple relevant disciplines (e.g. health, public services, social science, design, built environment). Searches were conducted by KH on 01.06.23, capturing literature from 01.06.12 and onwards, while any relevant literature on overcrowding before this date was carried forward from the previous Cochrane review (10).

Reference lists of more recent reviews captured were also checked, while tracing subsequent publication of data from any identified protocols and checking any ongoing reviews from PROSPERO. For relevant primary studies, we further investigated their reference lists and searched study names in Google Scholar to supplement with any ‘sibling’ records where needing information on contexts and/or mechanisms related to the interventions. Additional grey literature was searched such as the Healthcare Management Information Consortium (HMIC), in addition to websites of participating authorities and supplemented with relevant reports suggested by expert panel members (which were all checked for relevance using the below reported eligibility criteria).

Search terms included both subject heading and free text terms, informed by a previous WHO-review (9) on the health impacts of overcrowding, as well as overcrowding definitions and categories of interventions from expert panel discussions. From the staff expert panels we added a stronger focus on (home) environment and ‘congestion’ and health, while from the resident expert panels green or play space, communication from housing authorities and cultural strategies. The search strategy and combination of terms was developed in one database (Ovid MEDLINE) and amended as required for each database, with a sample of the strategy in Additional file 2.

Studies were then de-duplicated in Rayyan (Qatar Computing Research Institute) software, assessed on title and abstract by one reviewer (KH) (with another (EE) assessing 10% to discuss disagreements), while any potentially relevant studies underwent more detailed examination against the eligibility criteria on full-text by two independent reviewers (KH, EE). They resolved all discrepancies through discussion, but any unresolvable discrepancies would have been adjudicated by a third independent reviewer (JS).

### Eligibility criteria

#### Types of studies

Qualitative, quantitative and mixed-methods evaluations were included, with no restrictions on study design. Position papers, editorials or commentaries that did not report empirical results but that theorised (informally) about the relative effects of particular strategies were also included.

Publication status could be either peer-reviewed or grey literature such as conference papers, policy documents, project initiation documents (etc.). However, due to retrieval time and costs and the rapid, resource-limited nature of the review, we did not pursue any book chapters or academic theses records. For the same reason, all included records were also limited to the English language.

#### Domain/population

We initially prioritised relevant strategies affecting families, but in consultation with the expert panels we included HMOs (house in multiple occupation) where such information was available.

We included overcrowding interventions related to all physical house types that are static, inclusive of sheltered houses (but not residential care homes as it is unlikely that families or people in HMOs will be housed there). We excluded mobile homes such as house boats or caravans, which in general are more likely to serve recreational uses.

Although the focus was on developing recommendations specifically tailored to urban contexts in England, we also included evidence from comparable contexts (other OECD/partner countries (19)) that might provide transferable lessons (excluding less relevant contexts such as rehousing from slums in lower-income countries from the previous Cochrane review (10)).

#### Intervention/exposure

We included evaluations of strategies provided by an agency working at a local level with an explicitly stated aim (wholly or partially) to address housing overcrowding and/or where it had an effect on overcrowding or residents’ experience of it.

Interventions did not include a change of housing conditions due to other life events such as natural disasters. It also excluded *ad hoc* improvements, if outside of a housing programme addressing overcrowding, such as housing redesign or decorations initiated by householders themselves, minor repairs such as fixing of leaking pipes and broken windows, standard fire or injury prevention measures, modifications needed *irrespective* of overcrowding for e.g. mobility/medical reasons.

#### Comparator(s)/control

Assessment of outcomes before and after overcrowding interventions, comparable areas where certain interventions were not tried or different interventions were tried.

#### Outcomes

Any direct measures of health or mental and physical illness as well as self-reported well-being and quality of life. We also considered any impact upon overcrowding *per se*, i.e. changes to physical environment and/or residents’ perceptions/experiences of environment as an output (e.g. changes to occupancy levels, changes to space or use of space, changes to satisfaction with dwelling). We considered housing condition outcomes as relevant to health because there is extensive literature that has demonstrated the association between housing conditions and various health outcomes (11). Health service use was originally not considered an included outcome, but again due to the association with overcrowding and health conditions – which healthcare access may help alleviate (11) – it was also included. Moreover, we considered additional social and socio-economic outcomes if acting as potential determinants of health such as social inclusion/exclusion, education and employment measures. We include within this food insecurity as a potential determinant of health, as proposed by expert panels. Adverse effects of interventions were also included.

## Data extraction

A data extraction form was piloted and amended as necessary. Data extraction was completed by one reviewer (KH) in Excel software and checked by another (either a researcher (JS, EE) or for some local/regional documents expert panel members from these contexts checked). Data was extracted on key features within the review scope and on components considered important for a realist review such as study design, context (e.g. geographical setting, housing tenure, definition and level of overcrowding, populations affected/included), mechanisms/approach/aims and outcomes.

### Relevance and rigour assessments

Following realist review standards (13), the contribution of sections of data within a document was assessed based on two criteria: relevance and rigour. Relevance was determined based on the criterion of whether sections of text within a document were deemed to be relevant enough to contribute to theory testing and/or building (13), with particular attention to our inclusion/exclusion criteria outlined above. For realist approaches, judgement of rigour may not include an appraisal tool (13) – although in our endeavour to standardise assessments of ‘credibility’ and ‘trustworthiness’ of sources (13), where possible (i.e. for peer-reviewed literature) rigour was determined through the Mixed Methods Appraisal Tool (MMAT) (20) adapted to relevant study designs. This was used by two independent reviewers (KH, EE) with all discrepancies resolved through discussion, but if they had not been resolvable adjudicated by a third independent reviewer (JS). We tabulated all our final assessments as well as comment in the main text on whether there were any concerns based on the MMAT assessments related to individual studies. It is specifically worth noting that ‘incomplete outcome data’ was a common criterion with ambiguous interpretation and required agreement on a threshold (we operated with 80 percent completion rate as threshold).

### Data synthesis

In the protocol we pre-specified a consideration of meta-analyses, but recognised the little and heterogenous evidence available in previous reviews within housing to enable meta-analyses (10). Similarly, this was also the case within our more specific scope on overcrowding. As also in accordance with our protocol, we therefore proceeded to conduct a narrative synthesis (21) that examined emerging patterns around the contexts affecting potential mechanisms (as interventions, mediating factors and pathways) which in turn may lead to outcomes.

We grouped evaluated interventions from the peer-reviewed literature into categories of similar types of interventions. For each category, we constructed initial programme theories formed as CMO configurations, based on our reading of background literature and resident/policy maker engagement before the study took place. For each category, we started with peer-reviewed data to refine the CMO, using additional information from ‘sibling’ papers (see searches/screening) or the grey literature where needed, and as checked against local stakeholder consensus in the second ‘validation’ expert panels (see stakeholder involvement subsection above), to understand the ways in which specific mechanisms of interventions may be implementable in urban contexts in England and any barriers to their success. For each of the intervention categories, we have generated and display figures of the CMO configurations including our initial programme theories and how these were supported or nuanced by insights from the literature (peer-reviewed and grey) and expert panels.

Albeit we recognise that the sources of data to test and develop programme theories are of relatively different origins and, as such, in the main text we present the peer-reviewed literature, grey literature and expert panels in turn within each of the intervention categories. Essentially, keeping the narrative presentation distinct in this manner may facilitate a better understanding of how each information source contributed to the CMO configurations, as well as to highlight the limitations also within the peer-reviewed literature in this field (as key to building recommendations for the future).

Furthermore, as we combined quantitative and qualitative data particularly within the peer-reviewed literature, we were also informed by the approach to mixed-methods synthesis (22) on reporting a common format in which one type of data is ‘translated’ into the other. In our case, we ‘qualitised’ quantitative data, meaning we narratively report results through words (whether e.g. significant effects were shown or not with a p-value of 0.05 as significance threshold).

For the effect measures, we refer specifically to Table 1. We focus on effects from the peer-reviewed literature and not in the grey literature documents, because predominantly the latter consisted of reports of what local authorities had done without a corresponding evaluation evidencing the relative effects/impacts of outcomes as linked to specific interventions.

**Table 1:** Data extraction and summary of interventions (peer-reviewed literature)

| REHOUSING (n=7 interventions) |  |  |  |  |  |
| --- | --- | --- | --- | --- | --- |
| INTERVENTION<br>(LOCATION) | OVERCROWDING<br>DEFINITION (LEVEL) | AIMS/MECHANISMS | EVALUATION<br>(DURATION) | SAMPLE | OUTCOMES (in bold) & RESULTS |
| GoWell (Glasgow, Scotland) (31) | ‘Inadequately sized homes’ (7 interviewees reported living in overcrowded conditions – results connected to these residents) | Rehousing to nearby areas in newly built homes or recently refurbished to meet national standards | Qualitative interviews study (one year) | N=23 households<br>Age=N/A<br>Gender=N/A<br>Ethnicity=N/A<br>Tenure=Social housing | **‘No perceived improvements’**= e.g. moved from high-rise flat with major damp to cottage flat with more localised damp problem. **‘Perceived improvement in environment but not health’**= e.g. physical and psychosocial environment improved, but insufficient to alleviate longstanding anxiety & depression. **‘Perceived improvements to environments & health’**= e.g. friends visited children without feeling unsafe & garden space for physical activity (play, gardening) |
| New Home, New You (Plymouth, England) (41) | N/A (N/A) | Rehousing followed by behavioural intervention of ‘capability, opportunity and motivation’ consisting of:<br>(1) Education (information to improve capability);<br>(2) Persuasion (motivational interviewing);<br>(3) Incentivisation (e.g. fortnightly vegetable bag);<br>(4) Training (cooking lessons to improve capability);<br>(5) Enablement (access to resources/opportunities) | Mixed methods study (one year) | N=111 residents<br>Age= Mean: 36.63 years<br>Gender= Female: 68.5%<br>Ethnicity= White British: 92.8%<br>Tenure=Social housing | **Wilcoxon two-tailed test of HAY [How Are You?] quiz for health-related behaviours**= 12 months vs. baseline: Z= -5.563*<br>**Mean difference of the Short Warwick-Edinburgh Mental Well-being Scale**= 12 months vs. baseline: 1.22*<br>**Residents’ perspectives:** feeling that new dwellings released residents from overcrowding in previous accommodation |
| Scottish Housing Health and Regeneration Project (Glasgow, Scotland) (33, 42, 50, 52, 53) | Whether ‘rooms too small’ in residents’ view (N/A) | Rehoused into newly developed general-purpose socially-rented home let with accompanying improvements in indoor conditions such as greater warmth, eradication of damp & more space | Quantitative pre/post study (2004-2005) (42, 50, 52, 53) | N=731 residents<br>Age= Mean: 43.2 (intervention)<br>Gender= Female: 76.9%<br>Ethnicity= White: 97.9%<br>Tenure=Mainly social housing | **Percentage difference that agreed that ‘rooms too small’**= -10.8%*<br>**P-value of changes in common symptoms**= Gain vs. no gain/loss in dwelling space: 0.13<br>**P-value of changes in wheezing**= Gain vs. no gain/loss in dwelling space: 0.14 |
|  |  |  | Qualitative interview study (2007-2008) (33) | N=22 households<br>Age= Range: 30-70+ years<br>Gender= Female: 86.4%<br>Ethnicity=N/A<br>Tenure=Mainly social housing | **Households’ perspectives**= decrease of problems vs. previous accommodation, in addition to overcrowding including associated issues such as damp, surrounding anti-social behaviour & unsuitable conditions for health |
| UK Millennium Cohort Study (32) | More than two people per room (more than 10%) | Assessment of impact of residential moves | Quantitative longitudinal study (2001-2006) | N= 5,505 households<br>Age= Families with children (0-5)<br>Gender=N/A<br>Ethnicity=overcrowding associated with Black ethnicity<br>Tenure=All types | **Level of overcrowding**= 11.5% (pre) vs. 4.5% (post)*<br>**Neighbourhood poverty**= 1 in 5 families either moved to a poorer area or remained within 30% of poorest areas |

| INTERVENTION<br>(LOCATION) | OVERCROWDING<br>DEFINITION (LEVEL) | AIMS/MECHANISMS | EVALUATION<br>(DURATION) | SAMPLE | OUTCOMES (in bold) & RESULTS |
| --- | --- | --- | --- | --- | --- |
| **Section 8 Housing Voucher (across the USA) (37, 46-49)** | Greater than 1 persons per room (3.74%) (Kole), less than 1 room per person in household (average size of household = 4) (Wood), 'rooms 'cluttered' (N/A) (Schmidt), number of persons per room undefined (N/A) (Van Ryzin) | US federal rental assistance to help residents move from one unit to another of (in theory) better quality. It involves residents receiving a certificate or housing voucher from an administering agency to be able to afford rent in a privately owned apartment, with the intervention providing a monthly subsidy covering the difference between the cost of rent & housing utilities (approximately 30% of residents' income) & what they can afford to pay | Mixed methods study including RCT (18 'quarters') (48, 49) | **N**=8,573 households<br>**Age**= Mean: 30.7 years<br>**Gender**= Female: 91.8%<br>**Ethnicity**= Non-Hispanic White: 19.6%; Non-Hispanic Black: 49.8%; Hispanic: 21.4%<br>**Tenure**=Majority rent (56.3%) | **Mean crowding reduction** vs. control= -48%*<br>**Intent-to-Treat impact on level of overcrowding** (unemployed heads)= -0.055<br>**Households' perspectives**= more living space, allowed some women to escape unhealthy relationships, with associated stress reductions. Mixed results on child well-being |
|  |  |  | Quantitative (RCT) (1994-2002) (46) | **N**=3,537 residents<br>**Age**= Range: 12-19 years<br>**Gender**= Female: 50.1%<br>**Ethnicity**= African American: 62.8%; Hispanic: 30.0%<br>**Tenure**=Social & private housing | **Beta (standard error) of whether 'rooms cluttered' (in association with asthma)**=<br>Total sample: -0.091 (0.199);<br>Boys: -0.012 (0.282);<br>Girls= -0.197 (0.237) |
|  |  |  | Quantitative panel study (1997-2003) (37) | **N**=84,782 households<br>**Age**=N/A<br>**Gender**=N/A<br>**Ethnicity**=N/A<br>**Tenure**= 'Owned & rented' | **Coefficient (standard error) between voucher increase & people per room**=<br>Total sample: -0.0081 (0.0028)*;<br>'Overcrowded' = -0.0820 (0.0224)*;<br>'Severely overcrowded' = -0.1603 (0.2369) |
|  |  |  | Quantitative cross-sectional study (1996) (47) | **N**=102,003 households<br>**Age**= Mean: 38.7 years<br>**Gender**=N/A<br>**Ethnicity**= 'Foreign-born': 54%<br>**Tenure**=Private housing | **Mean Number of Persons Per Room**=<br>Raw mean: 0.68; adjusted mean: 0.63 (compared to 0.70 for all low-income renters) |
| **Norwegian Housing Allowance (39)** | Living in one-room flat or housing with lower number of rooms (excluding kitchen & bathroom) than persons (11.4%) | Welfare entitlement calculated based on a 'gap formula' of income & housing expenses mirroring regional variations in housing costs as well as variations in the cost of good standard housing depending on the size of the households | Quantitative controlled pre/post study (2009-2010) | **N**=93,154 households<br>**Age**= Mean (household head): 52.61 years<br>**Gender**=N/A<br>**Ethnicity**=N/A<br>**Tenure**=Social & private renting | **Marginal effects of moving probability**=<br>14.3% higher vs. baseline*<br>**NB**: 50.8% who move out of a crowded situation also move into crowdedness |
| **Spanish charity assistance (Caritas Diocesana Barcelona) (38, 40, 51)** | More than one person per room (excluding toilets) but including members of other families (56.7%) | Assisted by a social worker & could receive economic/social assistance for families with housing affordability problems &/or in substandard dwellings | Quantitative pre-post study (one year) | **N**=140 households<br>**Age**=Majority aged between 30 and 44 years<br>**Gender**=Majority women<br>**Ethnicity**= Foreign-born: 94.8%<br>**Tenure**=Private rental | **Level of overcrowding**=<br>58% (pre) vs. 42% (post)*<br>**Bivariate associations between changes in overcrowding level & hours of sleep**=<br>Improved: 32.4% vs. Equal or worse: 15.7%*<br>**NB**: No significant improvements in self-reported health; GHQ-36; migraine or frequent headaches; respiratory problems |

HOME IMPROVEMENTS (n=6 interventions)
| INTERVENTION<br>(LOCATION) | OVERCROWDING<br>DEFINITION (LEVEL) | AIMS/MECHANISMS | EVALUATION<br>(DURATION) | SAMPLE | OUTCOMES (in bold) & RESULTS |
| --- | --- | --- | --- | --- | --- |
| **GoWell (Glasgow, Scotland) (26)** | People per room (max=5, mean=0.85, standard deviation=0.47) | Impact of five housing improvements: external/structural; security; warmth; internal; unspecified | Quantitative cross-sectional study (2005-2007) | **N**=3,738 residents<br>**Age**=N/A<br>**Gender**=N/A<br>**Ethnicity**=N/A<br>**Tenure**=Social housing | **Relationship people per room & perceived housing quality + psychosocial benefits**=<br>Reportedly no effects (but overcrowding effects not isolated from model) |
| **Heat with Rent (Glasgow, Scotland) (34)** | N/A (21.7% overcrowded of initial sample of 254) | Installation in all rooms of controlled heating system responding to external temperature. Tenants paid a fixed sum incorporated into their rent. The scheme addressed both dampness and cold in dwellings & problems associated with budgeting & fuel poverty | Quantitative longitudinal study (one year) | **N**=132 residents<br>**Age**= 0-4 years: 36%<br>5-11 years: 44%<br>12-15 years: 20%<br>**Gender**=N/A<br>**Ethnicity**=N/A<br>**Tenure**=Social housing | **Level of overcrowding**=<br>Intervention: 23.6% (pre) vs. 23.6% (post);<br>Comparison: 15.6% (pre) vs. 18.2% (post)<br>**Would not use rooms due to damp**=<br>Intervention: 20.0% (pre) vs. 9.1% (post)*;<br>Comparison: 26.0% (pre) vs. 35.1% (post) |
| **Housing Sustainability, Self-help and Upgrading (Texas, US) (30)** | More than 2 persons/bedroom (N/A) | Title regularisation of informal housing, followed by self-help & formal market loans: 72% had major home improvements, 32% of remodelled one or more rooms, 26% & 25% with flooring & roofing improvements, respectively, & between 15% & 18% improvements to garden or parking area | Quantitative repeated cross-sectional study (2002-2011) | **N**=106<br>**Age**= Mean: 52.43 years<br>**Gender**= Female: 73%<br>**Ethnicity**= Mexican: 83%<br>**Tenure**= Private (albeit regularising informal) housing | **Level of overcrowding**=<br>17 % (pre) vs. 7% (post) |
| **Housing renovation project (Malmö, Sweden) (45)** | According to Swedish Statistical Agency corresponding to more than 2 inhabitants per bedroom (75%) | One neighbourhood affected by substandard housing & needed renovations (court-mandated partial repairs of kitchens & bathrooms), compared to neighbourhood not receiving similar renovations | Quantitative controlled pre/post study (2010-2012) | **N**=51 families with 127 children<br>**Age**= Mean: 7.4 (intervention)<br>**Gender**= Female: 47.8%<br>**Ethnicity**= Swedish-born: 80%<br>**Tenure**=Mainly social housing or subsidised rent | **Level of overcrowding**=<br>Intervention: 75% (pre) vs. 80% (post);<br>Comparison: 57% (pre) vs. 50% (post)<br>**Self-reported positive health of children**=<br>Intervention: 74% (pre) vs. 86% (post);<br>Comparison: 78% (pre) vs. 80% (post)* |
| **Housing retrofitting project (Porto, Portugal) (44)** | Usable space as measured in survey item "house has enough space" (N/A) | Main upgrades of buildings on roof with thermal insulation added, windows with full replacement & ventilation with addition of dedicated devices such as mechanical extraction & self-regulating inlets | Quantitative controlled pre/post study (one year) | **N**=82 residents<br>**Age**= Median: 57 years<br>**Gender**= Female: 52.2% (intervention)<br>**Ethnicity**=N/A<br>**Tenure**=Social housing | **Post-intervention agreement only with statement that 'house has enough space'**=<br>88.9% (intervention) vs. 79.2% (comparison) |
| **Optimisation Design for Interior Space (Changchun City, China) (24)** | Lack of indoor functional unit space as mainly concentrated in hall, kitchen, balcony & storage space (92%) | Partially open & overlap space to have more than one function, arranging furniture according to evolving needs & relationships; if area cannot be increased, insert partitions for use by multiple people while retaining privacy, adding corridor, door, window to create 'spatial loop' & feel same space from multiple perspectives (as if it was larger) | Theoretical (N/A) | **N**=100 households<br>**Age**=N/A<br>**Gender**=N/A<br>**Ethnicity**=N/A<br>**Tenure**= Private (small housing) | **Perception of space**=<br>Postulate that proposed design features increase sense of extension & space fluidity |

CO-ORDINATION WITH HEALTHCARE AND WIDER SERVICES (n=2 interventions)
| INTERVENTION<br>(LOCATION) | OVERCROWDING<br>DEFINITION (LEVEL) | AIMS/MECHANISMS | EVALUATION<br>(DURATION) | SAMPLE | OUTCOMES (in bold) & RESULTS |
| --- | --- | --- | --- | --- | --- |
| **Healthy Housing Programme (Auckland, New Zealand)** (23, 27-29, 35, 36) | New Zealand Census of Populations and Dwellings – bedrooms needed based on demographic composition (but house visits revealed that the tenancy data did not capture full extent of overcrowding & all households in catchment area therefore included) | Reduce risk of meningococcal disease & other conditions associated with crowding, broadened to other domains e.g. social importance of home, housekeeping skills, improving linkages & co-ordination with social & health services. Consisted of initial assessment visit by public health nurse, subsequent action plan developed, reviewed by community clinician & discussed with household members. Solutions ensured houses incorporated design elements critical to health. Consultation throughout process, with referral to health & social welfare agencies also facilitated | Quantitative case-control study (2001-2009) (35, 36) | **N**=9,736 residents<br>**Age**=weighted in favour of younger age groups<br>**Gender**=N/A<br>**Ethnicity**=almost exclusively Pacific Islanders<br>**Tenure**=Social housing | **Principal diagnosis of respiratory or infectious diseases (Hazard ratios, 95% CIs)**=<br>Age 0-4 years: 0.88 (0.74, 1.05);<br>Age 5-34 years: 0.73 (0.58, 0.91)*;<br>Age 35+ years: 1.31 (1.09, 1.56)* |
|  |  |  | Qualitative interview study (2000-2003) (23, 27-29) | **N**=30 households, 19 programme providers<br>**Age**=N/A<br>**Gender**=N/A<br>**Ethnicity**=Pacific Islands & Maori people<br>**Tenure**=Social housing | **Householders' perspectives**= feeling ownership & more control, more rooms as well as space to move facilitating harmonious interactions (e.g. less sibling rivalry) & better reported health as 'downstream effect'.<br>**Providers' perspectives**= enhanced study spaces for children means education was re-prioritised, accommodating cultural needs |
| **Well Homes (Wellington, New Zealand)** (25, 43) | Identified by assessor where average number of people sleeping in a room exceeded two (66.5% overcrowded in study population, compared to 5.1% in total New Zealand population) | Families referred to relevant outreach organisations meeting housing & health needs, who carried out home visits, supplied necessary items to help residents make best use of their space such as beds, heaters & draught-stoppers, while requesting landlords to make any repairs & improvements. Further assistance provided to register on wait-list for social housing, checked that the residents received sufficient welfare entitlements, other services e.g. ventilation & budgeting advice | Quantitative cross-sectional study (2015-2018) (43) | **N**=895 residents<br>**Age**= <5 years: 56.6%<br>5-14 years: 22.9%<br>15-28 years: 10.3%<br>**Gender**= Female: 55.8%<br>**Ethnicity**= Māori: 43.9%<br>Pacific people: 32.0%<br>**Tenure**= Private rental: 40.4%<br>Social housing: 47.3% | **Provision of bedding**= 96.1%<br>**Provision of beds**= 83.3%<br>**Delivered social housing relocation**= 11.1% of those with action attempted |
|  |  |  | Qualitative interview study (N/A) (25) | **N**=21 programme providers<br>**Age**=N/A<br>**Gender**=N/A<br>**Ethnicity**=N/A<br>**Tenure**=Mainly residents assisted in private rental | **Providers' perspectives**= residents placed on social housing register, while in the meantime provision of beds & bedding, advice about heating & sleeping arrangements, may be protective. However lack of social housing meant many could not be rehoused, or experienced long waiting times – providers often had to manage expectations rather than be able to promise residents anything |

\* = significant at p-value of 0.05; CIs = confidence intervals; GHQ = General Health Questionnaire; RCT = randomised controlled trial
**NB:** As most of the overcrowded samples from the literature were from lower socioeconomic backgrounds, the figure does not include socioeconomic characteristics

## Results

The PRISMA flow diagram of searches and screening is shown in Figure 1. After duplicates were removed, 8,558 records were screened on title and abstract, with 139 of those identified as potentially relevant and requiring full-text screening (four of those could still not be retrieved following contact with authors). 109 records were excluded from the full-text stage. The most common exclusion reason was ‘no overcrowding’ (n=58) – either that the households reported on were not overcrowded, or with insufficient information to determine this (reasons for full-text exclusion of each individual reference are provided in Additional file 3).

**Figure 1:**
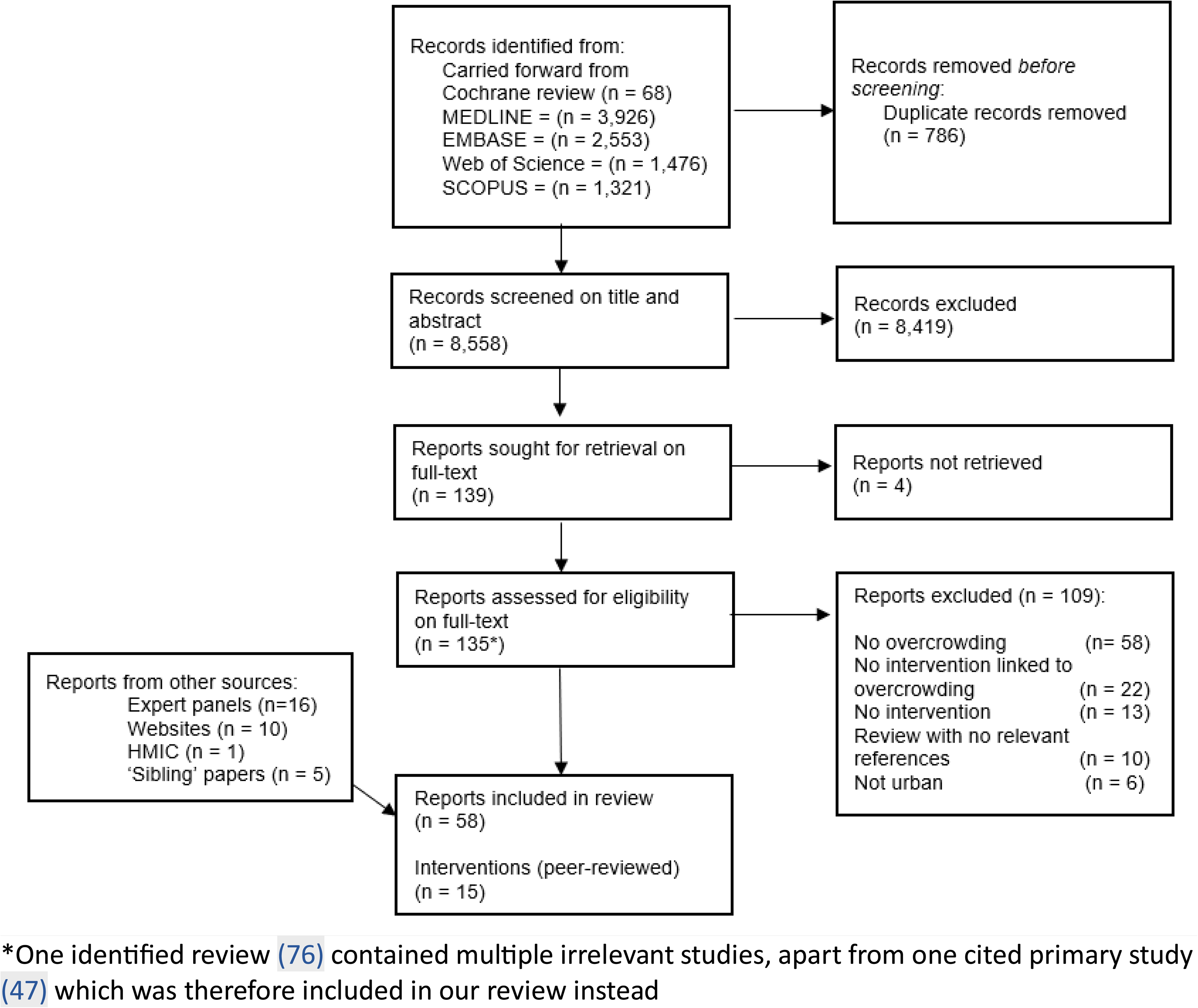
P**RISMA flow diagram of searches and screening**

Thirty-one peer-reviewed reports were included (26 from the initial searches (23–48) supplemented by 5 ‘sibling’ papers (49–53)), in addition to 27 grey literature documents (2, 5, 6, 8, 18, 54–75) related to the involved authorities (as either retrieved from the HMIC database (n=1), suggestions from expert panel members (n=16) or authorities’ official websites (n=10)). We centre our report below on the 15 evaluated interventions from the peer-reviewed literature^1^ – of which have also been supplemented by contextual information from the local authority grey literature and expert panels. The 15 interventions revolve around three categories: Rehousing (n=7 interventions (31–33, 37–42, 46–53); Home improvements (renovations/retrofitting) (n=6 interventions (24, 26, 30, 34, 44, 45)); Co-ordination with healthcare and wider services (home improvements as well as health/social care links) (n=2 interventions (23, 25, 27–29, 35, 36, 43)). More information on study characteristics and outcomes related to the 15 peer-reviewed interventions can be found in Table 1 (data extraction of information on context and mechanisms from the grey literature is also available from Additional file 4). Assessments on each MMAT criteria can be found in Additional file 5. To note that, informed by the first expert panels, we also searched but did not identify relevant literature on buy-back schemes, government promotions, or alleviation of overcrowding in HMO or hotel settings, as well as for health outcomes relating to COVID-19, the cost-of-living crisis or food insecurity.

### Rehousing

First, we tested/refined below the programme theory that support with rehousing (mechanisms) could benefit health (longer-term outcomes) for people whose household occupancy is over national standardised measures (e.g. bedroom standard), when it increases the probably of moving and increases experience/perceptions of space (proximal outcomes) (see Figure 2 for the CMO configuration).

**Figure 2:**
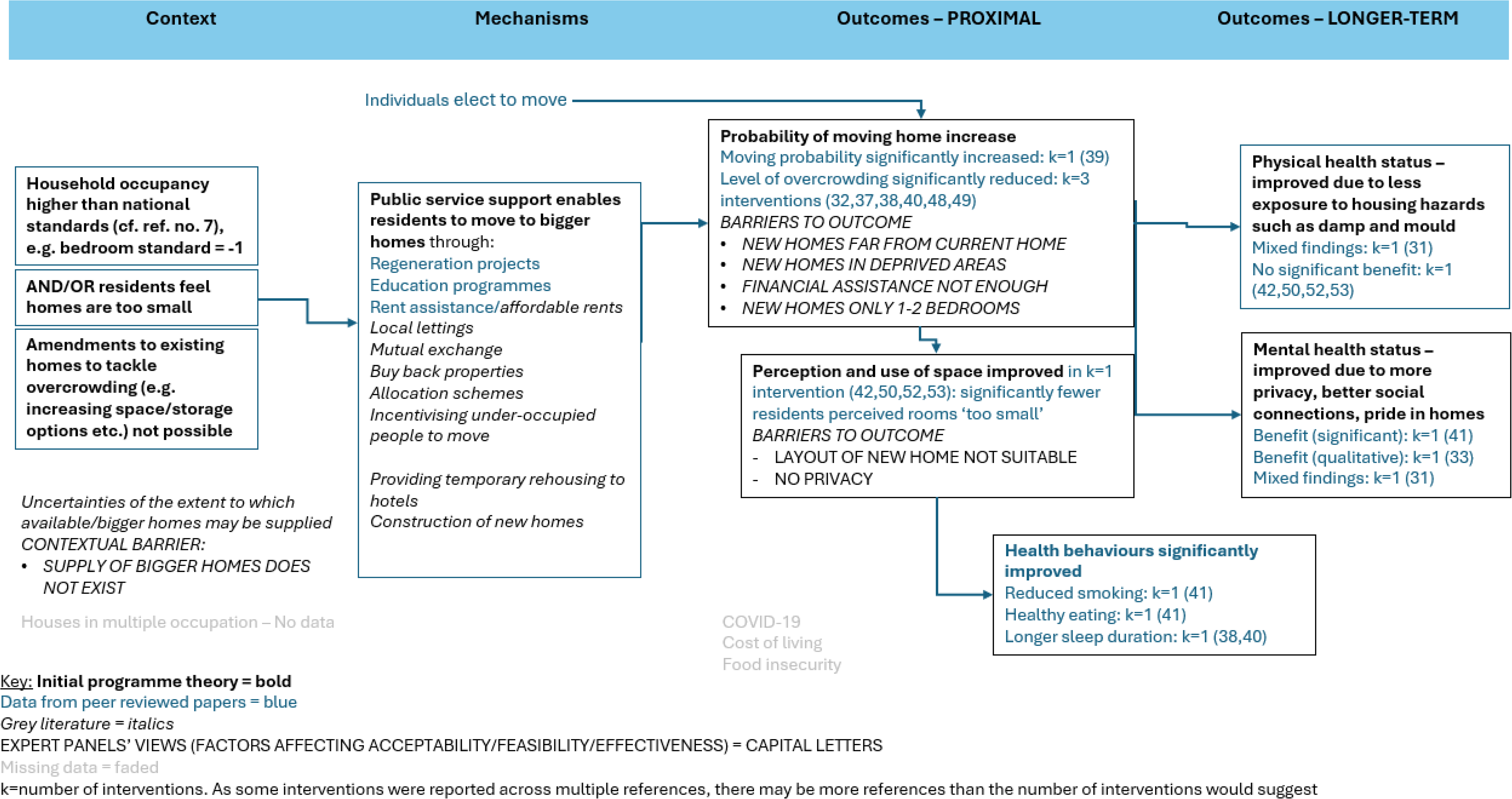
**Rehousing**

#### Peer-reviewed literature (n=7 interventions)

In the peer-reviewed literature, rehousing was mainly measured through interventions helping households to move out from overcrowded dwellings to (in theory) better quality dwellings (through regeneration projects (31, 33, 42, 50, 52, 53), additional educational/behavioural programme to integrate into the new setting and homes (41) and financial assistance to help with rent and housing costs (37–40, 46–49, 51)), but we also include a study of individual residential moves (32) due to its potential lessons for how moving house in general may alleviate overcrowding.

Four of the seven studies in this category are from the UK (31–33, 41, 42, 50, 52, 53). The level of overcrowding was reduced significantly (from 11.5% to 4.5%) in the UK Millennium Cohort Study (32), but it is unclear how many of these residential moves occurred due to councils helping residents to rehouse. In the Scottish Housing Health and Regeneration Project’s (SHARP) quantitative evaluation, there was a significant reduction in residents (by 10.8%) agreeing that their ‘rooms are too small’ (42). Incomplete outcome data was a concern; however, overcrowding was reportedly reduced also in the qualitative interview evaluation (33) for this project – with no identified quality concerns – in which residents reported more space both inside and outside including gardens. Another interview study (31) with no quality concerns from the Scottish context (GoWell) showed a mix of reported outcomes across three themes: ‘no perceived improvements’; ‘perceived improvement in environment but not health’; ‘perceived improvements to environments and health’ (see Table 1 for details). For further health outcomes, in a mixed methods study from Plymouth (England) (41) the quantitative evaluation showed significant improvements in mental well-being (mean difference= 1.22) and health behaviours such as healthy eating and reduced smoking (Wilcoxon two-tailed test, Z= −5.563) after one year in their new homes. A potential mechanism is that the intervention did not only consist of rehousing, but also of adequate follow-up of residents in their new dwellings with a behavioural programme of education and training to build skills to address housing issues. Such extensive follow-up and skills building appears relatively absent in the controlled pre-post evaluation (52) for the SHARP project, showing no significant health improvements.

Of the three remaining non-UK interventions, the most predominantly evaluated was US federal rental assistance (Section 8) to help residents move from one unit to another of (in theory) better quality (37, 46–49). In the randomised controlled trial (RCT) component of a mixed methods study (48) with no detectable quality concerns, level of overcrowding was significantly reduced (by 48%). In a panel study (37) with some concerns of representativeness and intervention administration, and a cross-sectional study (47) with no similar concerns, there was a decrease in the number of persons per room, but this was only significant in the panel study (37) for ‘overcrowded’ households (Coefficient (standard error)= −0.0820 (0.0224)) and not so for those previously in ‘severely overcrowded’ conditions. For health outcomes, less ‘cluttered’ conditions were experienced by children with asthma in an RCT (46) (albeit it failed to blind outcome assessors to the intervention and did not retain significance), while the other RCT (48, 49) showed mixed findings and no conclusive evidence for child well-being. The Norwegian welfare system, in contrast to the American, is not application-based and does not require ‘queueing up’ on a wait-list to receive the housing allowance, but rather everyone who is entitled to it will receive it similar to comparable European contexts (39). Although no *detectable* concerns, there was a lack of information to make up an assessment on three of the five MMAT items in a controlled pre-post evaluation focusing on mobility patterns (in contrast to home improvements that the housing allowance may also be used for) (39). It showed that the probability of moving homes significantly increased (by 14.3%), but that around half move into another situation of crowdedness rather than escaping it. Finally a non-governmental organisation in Barcelona, Spain (Caritas Diocesana) (38, 40, 51) provided economic assistance (as well as support from a social worker) for families in substandard dwellings (e.g. to escape overcrowding). The pre-post evaluation (38, 40), suffering somewhat from low sample size and loss to follow-up, showed significantly reduced overcrowding (by 16%) as well as longer sleep duration (32.4% improved vs. 15.7% equal or worse).

#### Grey literature

Out of the 27 included grey literature documents, 26 (2, 5, 6, 8, 18, 54–57, 59–75) provided information relevant for rehousing in England, helping us to understand the mechanisms by which public services have sought to help people to move (which was not as richly described in the peer-reviewed literature). This revealed schemes such as e.g. local letting opportunities and mutual exchanges to support residents to move (67), constructions of new homes (69) or buying back abandoned and poorly managed property from the Right-to-Buy scheme – which had allowed previous tenants to buy rented properties – to free these up for residents in need (61, 71, 75). However, space constraints, particularly in dense urban environments such as inner-city London, were highlighted (2). As such, rehousing within London was not always an option and either resulted in residents having to move elsewhere, being temporarily rehoused to hotel facilities, or waiting as long as decades on social housing registers (5). Allocation schemes did not always reward enough ‘points’ to households to be prioritised for housing allocation, as their overcrowding was not seen as ‘severe’ enough or considered in the same imminent need as e.g. homeless people (5). Alternatively, ‘affordable’ private rent schemes may not be within the price range of all households (56). Some other schemes were therefore set up to move residents for example to seaside and country homes (62), prioritising under-occupying residents in e.g. London that had more bedrooms available so that their move could free up sufficient space for overcrowded residents within the city.

### Expert panel validation

The staff panel concurred with caveats from the literature in reporting that proposed rehousing sites were often away from residents’ communities and only some had been willing to move to these.

Residents confirmed that more space in general was usually only available outside of London, with additional outdoor space being a specific need for children with neurological conditions such as autism. However, prospective rehousing sites were often in relatively socioeconomically deprived neighbourhoods and with a prospective break-up of their communities affecting both available support networks and their sense of belonging. A further concern was not only the location, but the actual dwellings they were rehoused to, with open plans offering insufficient room for privacy. Combined with this, residents were frustrated by the fact they had to accept the offer immediately without having a chance to see inside dwellings first. There was a lack of tailoring to households’ needs and inadequate accommodation for special needs prevented some households from moving in the first place. Furthermore, residents felt that new builds in London were designed to prioritise single professionals rather than families. In relation to any financial incentives to move or rent elsewhere, residents (as well as the grey literature) indicated that these are limited in scope or, if available, may be means-tested and risk compromising benefit caps. Also, residents experienced rental prices as so high, and especially in London, that any financial incentives might be insufficient.

### Home improvements

Our initial programme theory proposed that in overcrowded homes it is more likely that the space/layout/storage fails to meet the household’s needs and that quality issues affect dwellings including unusable rooms or damaged furniture (e.g. due to damp/mould damage, or other housing hazards such as rodent infestation) (context). Improvements (mechanisms) to space in residents’ current dwelling or their surrounding environments – either quantitatively in actual physical space or qualitatively as experienced amount of available space – could offset the need to move elsewhere (proximal outcomes) and improve health/well-being (longer-term outcomes) by making the home environment safer and use of space better (proximal outcomes) (see Figure 3 for the CMO configuration).

**Figure 3:**
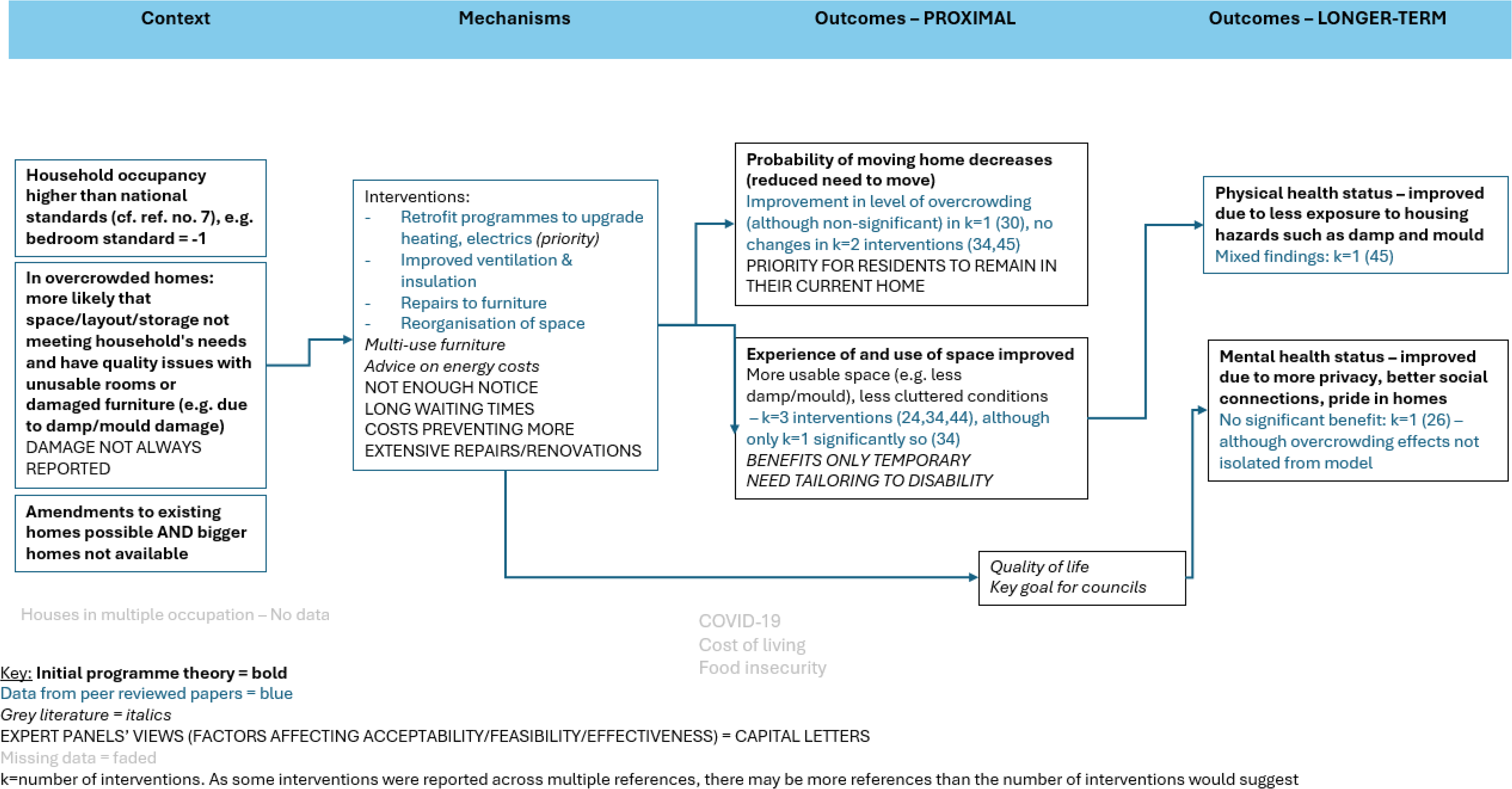
**Home improvements**

#### Peer-reviewed literature (n=6 interventions)

Two home improvement interventions related to retrofitting concentrating on upgrades to heating systems, ventilation, insulation, and electric efficiency to address functional issues such as dampness and mould (34, 44); two related to renovations or re-organisation of space, repairs to or addition of furniture to increase the qualitative amounts of usable space (24, 45); and two combined renovations and retrofitting for general home improvements with overcrowding outcomes (26, 30).

A longitudinal study from Scotland (34) retrofitted all rooms with a heating system with no significant differences in overcrowding, but a significant *reduction* (for almost 11% of residents) in rooms they were *not* able to use due to dampness (i.e. indicating some containment of this issue to free up available space). Based on available information, the study fulfilled all MMAT criteria apart from incomplete outcome data. A controlled pre-post evaluation from Portugal (44) of thermal insulation (roof), full replacement of windows and improved ventilation evidenced a non-significant increase in the perception that the ‘house has enough space’, but only one MMAT criterion was fulfilled.

For renovations, a controlled pre-post evaluation from Sweden (45) of only partial improvements to kitchens and bathrooms showed no significant differences in the level of overcrowding, while self-reported health of children showed signs of improvements in both the intervention and comparison area. However, the study did not fulfil any MMAT criteria. A theoretical postulation from China (24) – not fulfilling any MMAT criteria albeit with key information missing to answer most – illustrated how design features can increase the sense of extension and fluidity of space to combat the experience of overcrowding. This could be important as children of different genders grow older, where e.g. a bathroom split up into sections (shower, sink, toilet) facilitates separation of intimate spaces.

A cross-sectional study from Scotland (26) combined renovation and retrofitting measures with reportedly no effects on overcrowding – albeit in this study less than half of the sample had received the home improvements. A repeated cross-sectional study over almost ten years of primarily Mexican migrants in USA (30), also included a mixture of renovations (remodelling of rooms, improvements to the garden) and retrofitting (floor and roofing repairs to retain warmth). It demonstrated a non-significant reduction in overcrowding (although outcome data was incomplete).

#### Grey literature

Ten (2, 5, 6, 54, 60, 65, 66, 68–70) out of the 27 included grey literature documents provided information on or revealed similar home improvement initiatives in England as those evaluated in the peer-reviewed literature. For example, for retrofitting local authorities recognised that quality of life depends on ventilation and heating (65), while measures to combat damp and mould included informal advice to manage energy costs (69). Renovation strategies included funding for space-saving furniture or multi-use home adaptations (e.g. to alleviate shared sleeping arrangements) (69).

Trialling of innovative architectural practices was indicated (65), with for instance moveable walls to roll rooms like the kitchen forward when needed and then back into space when not needed.

#### Expert panel validation

Both staff and residents expressed an interest in designs to generate homes that could be adapted as residents aged and developed according to changing needs, so that families did not have to keep moving. Staff also considered planning and implementing such solutions when designing new builds, rather than retrospectively as ‘emergency solutions’. Despite such aspirations, it was expressed that local funding is an issue, so that smaller or traditional initiatives of retrofit and renovations that are already available, might be prioritised.

Home improvements were valued as potentially enabling residents to stay – the preferred option for most residents rather than having to move – with home improvements alleviating some of the worst impacts of overcrowding. Yet, an overriding feeling amongst residents was that although such interventions themselves were not the issue, how they were delivered, or not delivered, was a concern. Residents expressed that waiting times may be long, or conversely that they are not given enough notice for inspections. Residents may refrain from asking for improvements in the first place, as they may be anxious that they will be wrongly accused of being responsible for any issues. Consistent with findings from the literature, although minor repairs might help in the short term, they are only a temporary solution if e.g. damp/mould keeps coming back. Some home improvements failed to be adequately tailored, for example disabled residents needing specially designed toilets.

### Co-ordination with healthcare and wider services

Our initial programme theory was that the health of residents living in overcrowding may be affected (context). Residents’ well-being may be improved (longer-term outcome) by better access to healthcare (mechanism) (see Figure 4 for the CMO configuration).

**Figure 4:**
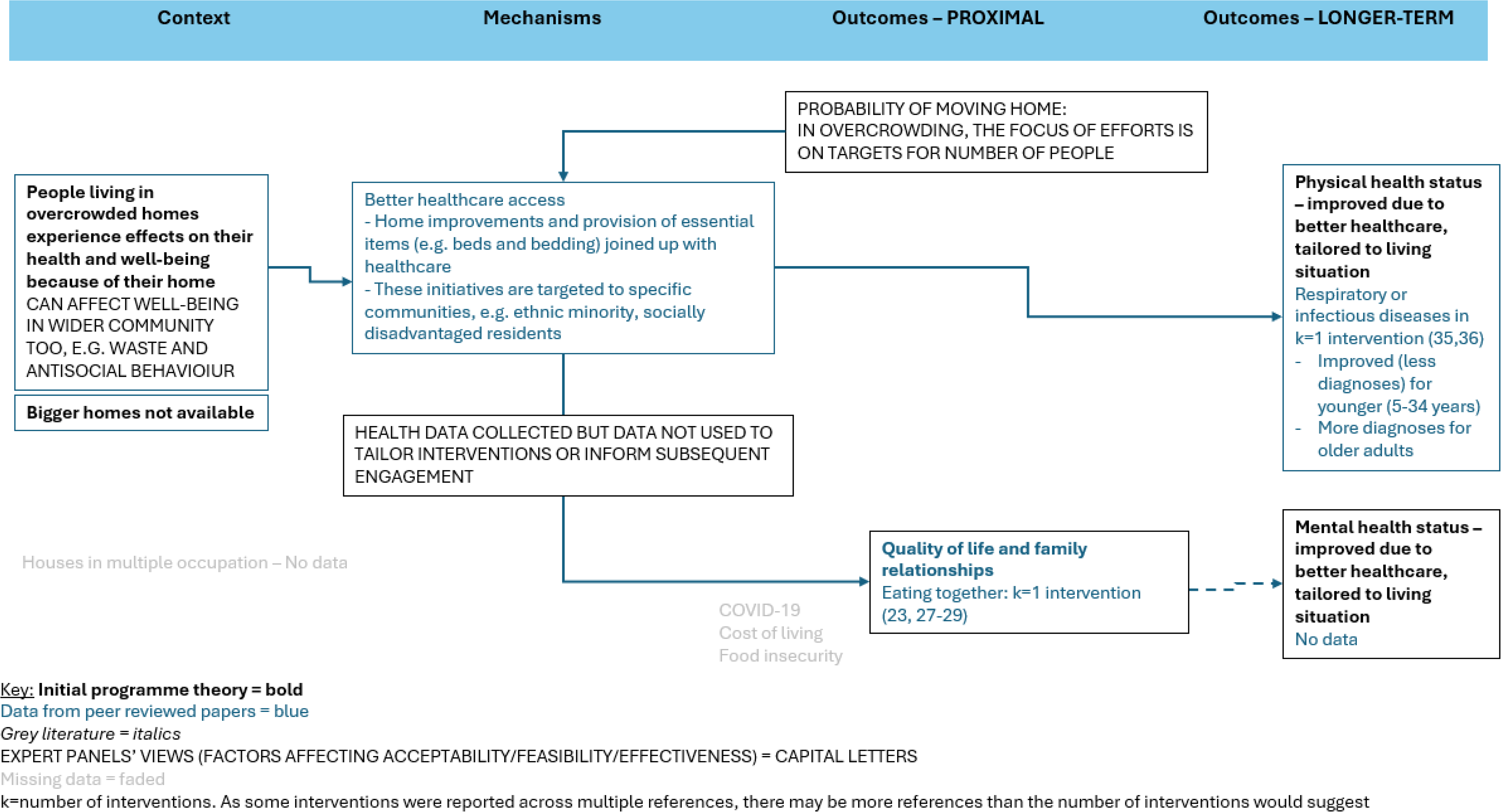
**Co-ordination with healthcare and wider services**

#### Peer-reviewed literature (n=2 interventions)

Positive results were indicated by two programmes from New Zealand (23, 25, 27–29, 35, 36, 43) when home improvements were joined up with healthcare particularly for ethnic minority and socioeconomically disadvantaged residents – providing access while recognising wider determinants of health than measures within existing properties alone.

The longest evaluation between 2001-2009 of the programme in Auckland (35, 36) – with no clear quality concerns – considered principal diagnosis of acute respiratory or infectious diseases including where “a strong causal link between the housing intervention and the illness could be postulated through reducing overcrowding” (p. 589) (36). The evaluation had mixed results depending on age. It significantly increased diagnoses for those aged 35 years or over, albeit the programme aimed at improving conditions particularly for younger groups and had a significant reduction for those aged 5-34 years (Hazard Ratio (95% confidence intervals) = 0.73 (0.58, 0.91)) and a non-significant reduction for those aged 0-4 years. Qualitative interviews with no quality concerns (23, 27–29), also suggested better health overall, and stronger family connectedness such as eating dinner together.

A quantitative evaluation of the programme in Wellington (43) showed efficient identification and delivery of items such as beds and beddings (although had incomplete outcome data), while residents in qualitative interviews (25) with no detectable quality concerns expressed how such items temporarily alleviated overcrowding when waiting on social housing registers.

#### Grey literature

Eleven (5, 18, 54, 55, 57–59, 66, 68, 71, 73) of the 27 included grey literature documents incorporated some recognition of the association between quality of housing and health, including corresponding measures to mitigate adverse health impacts. This could e.g. be through acknowledging the health concerns related to more severe levels of overcrowding (not only with a focus on the household *per se*, but also around the wider community in terms of the potential accumulation of waste and anti-social behaviour (58)), as well as overcrowding definitions that prioritised residents with certain diagnoses in councils’ allocation policies (55), or in more integrated ways with the role of healthcare highlighted in strategic plans for the future (71, 73).

#### Expert panel validation

Some staff noted they had a mandated task to reduce the number of people officially categorised as ‘overcrowded’ in their local authorities. This meant that initiatives that peer-reviewed literature suggest could improve health or well-being might not be considered if they did not change numbers living in overcrowding.

Residents expressed that councils might be aware of the impact on health. A lot of screening measures prevailed including councils collecting and using data for health and safety, according to residents, without listening to or following sufficiently up with them. Further, residents felt that currently they are not joined up sufficiently with healthcare services as they are either unsure of how systems work or may give up anyway as they felt they cannot book an appointment with the doctor when needed.

## Discussion

### Main findings

We conducted an RRR including a search for peer-reviewed and grey literature combined with resident and staff involvement, which allowed us to test our initial programme theories and identify promising interventions and mechanisms to alleviate the negative effects of overcrowding on households’ well-being. Consistent with our programme theory for this intervention and findings in peer-reviewed literature, rehousing (mechanism) was not always considered as ideal by residents (proximal outcome) who described a trade-off between more space vs. loss of social networks and potentially poorer quality housing or environment (longer-term outcomes). Also consistent with another programme theory and the literature, resident expert panels verified how in the case of home improvements (mechanisms), these may alleviate the worst impacts of overcrowding (outcomes) and address changing needs (context) especially if delivered from a design and planning stage (mechanism). Additionally, interventions to improve healthcare co-ordination and access (mechanisms) can be effective (outcome) and may be particularly appropriate for residents living for several years in overcrowded conditions (context).

### Comparison with previous research

This review updates the evidence base of the 2013 Cochrane review on housing interventions (10), providing a stronger focus on overcrowding. In the Cochrane review many interventions of potential relevance to overcrowding focused on rehousing. This left some questions unaddressed about what can be done when residents cannot move. Our review adds to this while providing a broader overview including alternative mechanisms that may be available for councils to reduce overcrowding or alleviate its negative health impacts. We acknowledge that a more recent and full realist review has been published on the topic of holistic housing renovations (77), but this concerns adults in disadvantaged neighbourhoods more broadly than the specific issue of overcrowding. In fact, of the nine pathways to improved health that those authors presented, only one mentioned addressing overcrowding as a subset of multiple actions to support the particular pathway of physical housing improvements combined with health referrals – and, as such, gives further validation to our key finding on healthcare co-ordination. Another relatively recent review (although not systematic or realist) (78) is also of relevance to the findings on how improved layout and space can have a positive impact on overcrowded children, in which this may provide private space to be alone and serve a protective well-being effect enabling children to regulate negative stimuli in the house due to overcrowding, such as stressful social interactions or noise. The importance of the home for children cannot in this sense be overestimated, with other literature highlighting that children often have less of a world outside the home than adults and may need private space to concentrate on schoolwork (79, 80).

### Limitations

There are some limitations. Firstly, time and resource constraints necessitated a ‘rapid’ review format. As this resulted in a shorter window for iteration or the possibility of adding further documents, the present review may not be as comprehensive as a *full* realist review. However, we conducted a comprehensive search in electronic databases similar to systematic review standards and benefited from key stakeholders pointing us to potentially missing literature, as well as further searching the grey literature on participating authorities’ websites. ‘Sibling’ papers were then identified to provide any additional information on context and/or mechanisms related to interventions evaluated in the peer-reviewed literature. Another caveat is that some care should be taken in generalising to urban contexts in England in general. For example, practical considerations and the need to facilitate in-person events to enhance participation, necessitated restriction of the residents’ expert panels to two councils within London. We do believe though that the expert panels and particularly accounting for residents’ perspectives including of seldom heard voices of ethnic minority residents, may be considered a strength. Finally, although the grey literature directed us to interventions implemented in participating contexts, and may suggest more evidence can be found in the grey compared to academic literature, these were typically not evaluated or as robustly evaluated as the peer-reviewed literature. Hence, the ways in which these have an impact or not, may not invariably be as certain despite the expert consultations and comparisons with similar evaluated interventions from the peer-reviewed (and occasionally international) literature.

### Recommendations for research

A realist review format was deemed necessary, with studies lacking specific outcome measures for overcrowding and using multiple study designs of variable quality and across contexts. As such, the present review benefitted from complementary information from the grey literature and stakeholder groups – enhancing local relevance and the prospect of achieving intended impacts of interventions.

In light of this observation, we offer four research recommendations. Firstly, more research is needed into interventions that are not concerned with rehousing only, but also other alternatives that allow residents to stay in their current homes such as home improvements. Secondly, evaluations should incorporate consideration of both the intervention itself as well as its implementation, from residents’ perspectives. This will help focus on a wider set of outcomes of importance to residents themselves and their qualitative amount of usable space, rather than merely through housing registers/metrics quantifying the numbers of people per rooms. Thirdly, findings need to be disaggregated by population groups. There were only a few examples of this, such as e.g. where home improvements combined with health and social care links appeared to have a stronger preventative effect in younger age groups with less prior exposure to overcrowding, when compared to older household members. Primary studies should therefore improve assessment of outcomes across multiple sociodemographic characteristics such as age and gender (etc.) and do so consistently across all intervention categories for comparison. Finally, the lack of evidence on some intervention categories suggested by the expert panels on buy-back schemes, government promotions, or alleviation of overcrowding in HMO or hotel settings, as well as overall measurement of health outcomes across any intervention categories relating to recently prominent issues in the UK such as the COVID-19 pandemic, cost-of-living crisis or food insecurity, should also be explored.

### Recommendations for policy/practice

This review focused on interventions that could be implemented at a local level in England. Our findings however also clearly demonstrate that local policy needs to be supported by national policy and needs to take into account local and national context. For example, staff described how councils need to balance limited resources to tackle overcrowding alongside other housing priorities such as shelter for a growing homeless population (61), which may be tackled through national prioritisation of affordable housing supply (e.g. building of social housing) (81). However, a lack of or reduced funding over many years from central and more regional structures (68) have potentially worsened the ‘housing crisis’. The accumulating challenges of post-pandemic recession, unemployment, increased living costs and rents, lack of discretionary housing payments and inappropriate benefit caps for those in need, as well as unfair evictions from the private rented market (54), speak to the larger concerted effort needed to tackle broader socio-economic inequalities – probably far beyond investing in effective and relevant overcrowding measures alone.

There are of course solutions available to councils in the immediate term – while realising that to be effective across urban contexts and population groups they may require further standardisation across councils through nationwide campaigns. In particular we offer four recommendations for policy and practice. Firstly, rather than just focusing on the prevalence of overcrowding in the prioritisation of policy, a more explicit consideration can be made of health and well-being. This is because the prevalence of overcrowding is not likely to drop immediately and there may be a transitional period in which our review has suggested solutions such as various home improvement initiatives that may alleviate the worst impacts of overcrowding and improve family health and well-being whilst still being in overcrowded conditions. Secondly, overcrowding should be considered as a council-wide issue that may not be tackled within the housing sector alone. Thirdly, the grey literature revealed that some of the evaluated interventions and mechanisms from the peer-reviewed literature are in place within local authorities – as such it may not be necessary to ‘re-invent the wheel’ completely, but ensure these are more closely aligned to residents’ needs (for example longer time to prepare for inspections or selection/move to any rehousing opportunities).

This may not be a separate endeavour for authorities, but in fact it is suggested that for initiatives to work, better signposting to organisations that support residents in their current or new environments may be needed. This then relates to the final policy/practice recommendation of more accurate and ongoing communication, such as regular status updates on residents’ applications and available options to alleviate overcrowding in the immediate as well as the longer-term. Ensuring that residents experience communication and any messages as appropriate may necessitate further co-design of engagement campaigns with the affected communities themselves. As the evidence shows that ethnic minority people are disproportionately affected by overcrowding in urban contexts in England (6), it is also pivotal that potential language barriers are addressed and sufficient translation services provided for non-English languages widely spoken within local communities.

## Conclusions

Reducing the prevalence of overcrowding requires national level and long-term policy changes to increase the supply of affordable homes. Therefore, rehousing will not be a feasible solution in the short term for many residents living in overcrowded homes. Moreover, this review found that rehousing is not always the optimal solution for the well-being of residents in overcrowded homes. However, it provides evidence of how other interventions such as home improvements and co-ordination with healthcare could address well-being when residents in overcrowded accommodation cannot or do not wish to move. Although our focus was on making recommendations for urban contexts in England, we have also included international peer-reviewed literature and our conclusions may be transferable to comparable contexts affected by household overcrowding.

## Supporting information

Additional file 1 - List of items to be included when reporting a realist synthesis

Additional file 2 - Sample search strategy

Additional file 3 - List of excluded references & reasons on full-text

Additional file 4 - Data extraction of grey literature

Additional file 5 - MMAT assessments

## List of abbreviations

CMO: contexts/mechanisms/outcomes
HMIC: Healthcare Management Information Consortium
HMO: house in multiple occupation
MMAT: Mixed Methods Appraisal Tool
RAMESES: Realist And MEta-narrative Evidence Syntheses: Evolving Standards
RCT: randomised controlled trial
RRR: rapid realist review
SHARP: Scottish Housing Health and Regeneration Project
WHO: World Health Organization

## Declarations

### Ethics approval and consent to participate

Panel members were not considered research participants, but experts in the review with ethical approval not required. However, they were provided with full details including an information sheet on the purpose of the review and panels and they could withdraw at any time without providing any reason.

### Consent for publication

Not applicable. Permission was requested for panels to be audio-recorded to enable researchers to focus on discussion, which in one case was denied, but permission was given to take notes. However, recordings and notes of panels were not subject to analysis of individual responses and we do not report on individualised experiences or sentiments, but only on views for which consensus was indicated by multiple voices.

### Availability of data and materials

Data analysed during this study are included in this published article [and its Additional files].

## Authors’ contributions

JS and MU developed the idea for the overall project, KH the review protocol. KH designed and conducted the searches, which were further refined through suggested additions by JS, MU and the expert panels. KH and EE screened records for inclusion. KH and RMM extracted relevant data, checked by JS or EE (with some staff expert panel members also involved in data extraction related to their local authorities). KH and EE performed quality assessment, with differences to be reconciled by JS. KH conducted the narrative synthesis presented for the expert panels’ consideration. MO and PS recruited participants for the Tower Hamlets residents’ expert panels, JS and MU for the staff’s expert panels. They also facilitated the respective expert panel sessions with KH. KH drafted the manuscript of the paper. All authors contributed to commenting on drafts, suggesting revisions of the manuscript and approved the final version.

## Data Availability

Data analysed during this study are included in this published article [and its Additional files].

## Acknowledgements

We would like to thank all participants that participated in the expert panels. We would also like to give a special thanks to Salma Begum and Majida Sayam and their organisation Jannaty Women’s Social Society (225-229 Seven Sisters Rd, London, N4 2DA, charity number: 1151143) who recruited participants and hosted (and facilitated with KH) the Islington residents’ expert panels. As well as to Dr Lorraine McDonagh for taking time to look over a draft of the manuscript and providing helpful feedback.

## Footnotes

1 The lower number of interventions than reports is because for some interventions, multiple papers reported on different aspects of the same intervention.

