## Additional file 1 - List of items to be included when reporting a realist synthesis for "Interventions that could mitigate the adverse effects of household overcrowding: A rapid realist review with stakeholder participation from urban contexts in England"

| **List of items to be included when reporting a realist synthesis (from Wong et al 2013)**  **Additional file 1**  TITLE | | | **Reported on page:** |
| --- | --- | --- | --- |
| 1 |  | In the title, identify the document as a realist synthesis or review | 1 |
| ABSTRACT | | |  |
| 2 |  | While acknowledging publication requirements and house style, abstracts should ideally contain brief details of: the study's background, review question or objectives; search strategy; methods of selection, appraisal, analysis and synthesis of sources; main results; and implications for practice. | 2-3 |
| INTRODUCTION | | |  |
| 3 | Rationale for review | Explain why the review is needed and what it is likely to contribute to existing understanding of the topic area. | 3-6 |
| 4 | Objectives and focus of review | State the objective(s) of the review and/or the review question(s). Define and provide a rationale for the focus of the review. | 4-6 |
| METHODS | | |  |
| 5 | Changes in the review process | Any changes made to the review process that was initially planned should be briefly described and justified. | 9, 12 |
| 6 | Rationale for using realist synthesis | Explain why realist synthesis was considered the most appropriate method to use. | 5-6 |
| 7 | Scoping the literature | Describe and justify the initial process of exploratory scoping of the literature. | 9 |
| 8 | Searching processes | While considering specific requirements of the journal or other publication outlet, state and provide a rationale for how the iterative searching was done. Provide details on all the sources accessed for information in the review. Where searching in electronic databases has taken place, the details should include, for example, name of database, search terms, dates of coverage and date last searched. If individuals familiar with the relevant literature and/or topic area were contacted, indicate how they were identified and selected. | 9-10 |
| 9 | Selection and appraisal of documents | Explain how judgements were made about including and excluding data from documents, and justify these. | 10 |
| 10 | Data extraction | Describe and explain which data or information were extracted from the included documents and justify this selection. | 12 |
| 11 | Analysis and synthesis processes | Describe the analysis and synthesis processes in detail. This section should include information on the constructs analyzed and describe the analytic process. | 13-14 |
| RESULTS | | |  |
| 12 | Document flow diagram | Provide details on the number of documents assessed for eligibility and included in the review with reasons for exclusion at each stage as well as an indication of their source of origin (for example, from searching databases, reference lists and so on). You may consider using the example templates (which are likely to need modification to suit the data) that are provided. | 14-15 and Figure 1 |
| 13 | Document characteristics | Provide information on the characteristics of the documents included in the review. | 15 and Table 1 |
| 14 | Main findings | Present the key findings with a specific focus on theory building and testing. | 16-28 including Figures 2, 3 and 4 |
| DISCUSSION | | |  |
| 15 | Summary of findings | Summarize the main findings, taking into account the review's objective(s), research question(s), focus and intended audience(s). | 28 |
| 16 | Strengths, limitations and future research directions | Discuss both the strengths of the review and its limitations. These should include (but need not be restricted to) (a) consideration of all the steps in the review process and (b) comment on the overall strength of evidence supporting the explanatory insights which emerged.  The limitations identified may point to areas where further work is needed. | 29-31 |
| 17 | Comparison with existing literature | Where applicable, compare and contrast the review's findings with the existing literature (for example, other reviews) on the same topic. | 28-29 |
| 18 | Conclusion and recommendations | List the main implications of the findings and place these in the context of other relevant literature. If appropriate, offer recommendations for policy and practice. | 31-33 |
| 19 | Funding | Provide details of funding source (if any) for the review, the role played by the funder (if any) and any conflicts of interests of the reviewers. | 35 |

**Reference:** Wong G, Greenhalgh T, Westhorp G, Buckingham J, Pawson R. RAMESES publication standards: realist syntheses. BMC medicine. 2013;11:1-14.
