## Additional file 2 - Sample search strategy for "Interventions that could mitigate the adverse effects of household overcrowding: A rapid realist review with stakeholder participation from urban contexts in England"

### Additional file 2: Sample search strategy – Ovid MEDLINE

1.	Crowding/ or Housing/ or "Public Housing"/ or exp "Home Environment"/

2.	((propert* or hous* or home* or dwelling* or residenc* or occupanc* or accommodation* or flat* or apartment* or hotel* or motel* or "B and B*" or "B n B*" or BandB* or BnB* or "bed and breakfast*" or HMO* or "house in multiple occupation*" or "house of multiple occupanc*") adj3 (crowd* or overcrowd* or cramped or crammed or condition* or standard* or quality or space* or health* or environment* or congest*)).ti,ab.

3.	1 or 2

4.	"Program Evaluation"/

5.	((exten* or provi* or increas* or improv* or convert* or reconvert* or regen* or renovat* or retrofit* or "retro fit*" or repair* or rebuild* or intervention* or program*) adj2 (propert* or spac* or area* or hous* or home* or dwelling* or residenc* or occupanc* or accommodation* or flat* or apartment*)).ti,ab.

6.	(rehous* or "re hous*").ti,ab.

7.	((propert* or hous* or home* or dwelling* or residenc* or occupanc* or accommodation* or flat* or apartment* or mutual or incentive* or reward* or payment* or paid) adj3 (swap* or transfer* or exchange*)).ti,ab.

8.	((propert* or hous* or home* or dwelling* or residenc* or occupanc* or accommodation* or flat* or apartment*) adj3 (refurbish* or insulat* or ventilat*)).ti,ab.

9.	((priorit* or repriorit*) adj3 (list* or register* or scheme*)).ti,ab.

10.	((green* or play or commun* or storage*) adj3 (spac* or area*)).ti,ab.

11.	((council* or housing or charit*) adj3 (communicat* or update* or support* or guid* or advice or advis* or inform* or direct* or signpost* or translat* or language*)).ti,ab.

12.	(cultur* adj3 (intervention* or program* or strateg* or service*)).ti,ab.

13.	or/4-12

14.	3 and 13

15.	homeless*.ti,ab.

16.	exp homeless persons/

17.	15 or 16

18.	14 not 17

19.	limit 18 to (english language and humans)

20.	limit 19 to dt=20120601-20230601

21.	remove duplicates from 20
