## Additional file 5 - MMAT assessments for "Interventions that could mitigate the adverse effects of household overcrowding: A rapid realist review with stakeholder participation from urban contexts in England"

| **STUDY DESIGN 🡪** | **1. QUALITATIVE** | | | | | **2. QUANTITATIVE (RCT)** | | | | | **3. QUANTITATIVE**  **(NON-RANDOMISED)** | | | | | **4. QUANTITATIVE (DESCRIPTIVE)** | | | | | **5. MIXED METHODS** | | | | |
| --- | --- | --- | --- | --- | --- | --- | --- | --- | --- | --- | --- | --- | --- | --- | --- | --- | --- | --- | --- | --- | --- | --- | --- | --- | --- |
| **MMAT CRITERIA 🡪** | **1.1** | **1.2** | **1.3** | **1.4** | **1.5** | **2.1** | **2.2** | **2.3** | **2.4** | **2.5** | **3.1** | **3.2** | **3.3** | **3.4** | **3.5** | **4.1** | **4.2** | **4.3** | **4.4** | **4.5** | **5.1** | **5.2** | **5.3** | **5.4** | **5.5** |
| **HOME IMPROVEMENTS** | | | | | | | | | | | | | | | | | | | | | | | | | |
| GoWell (Scotland) (1) |  |  |  |  |  |  |  |  |  |  | Yes | Yes | N/a | Yes | No |  |  |  |  |  |  |  |  |  |  |
| Heat with Rent (Scotland) (2) |  |  |  |  |  |  |  |  |  |  | Yes | Yes | No | N/a | Yes |  |  |  |  |  |  |  |  |  |  |
| Housing Sustainability, Self-help and Upgrading (Texas, US) (3) |  |  |  |  |  |  |  |  |  |  | Yes | Yes | No | N/a | N/a |  |  |  |  |  |  |  |  |  |  |
| Housing renovation (Swe.) (4) |  |  |  |  |  |  |  |  |  |  | N/a | No | No | No | No |  |  |  |  |  |  |  |  |  |  |
| Housing retrofitting (Port.) (5) |  |  |  |  |  |  |  |  |  |  | N/a | Yes | No | No | No |  |  |  |  |  |  |  |  |  |  |
| Optimisation Design (China) (6) |  |  |  |  |  |  |  |  |  |  |  |  |  |  |  | N/a | No | No | N/a | N/a |  |  |  |  |  |
| **DETERMINANTS OF HEALTH** | | | | | | | | | | | | | | | | | | | | | | | | | |
| Healthy Housing (quantitative) (7, 8) |  |  |  |  |  |  |  |  |  |  | Yes | Yes | Yes | Yes | N/a |  |  |  |  |  |  |  |  |  |  |
| Healthy Housing (qualitative) (9-12) | Yes | Yes | Yes | Yes | Yes |  |  |  |  |  |  |  |  |  |  |  |  |  |  |  |  |  |  |  |  |
| Well Homes (quantitative) (13) |  |  |  |  |  |  |  |  |  |  |  |  |  |  |  | N/a | Yes | No | Yes | Yes |  |  |  |  |  |
| Well Homes (qualitative) (14) | Yes | Yes | Yes | Yes | Yes |  |  |  |  |  |  |  |  |  |  |  |  |  |  |  |  |  |  |  |  |
| **RE-HOUSING** | | | | | | | | | | | | | | | | | | | | | | | | | |
| GoWell (Scotland) (15) | Yes | Yes | Yes | Yes | Yes |  |  |  |  |  |  |  |  |  |  |  |  |  |  |  |  |  |  |  |  |
| New Home, New You (Eng.) (16) |  |  |  |  |  |  |  |  |  |  |  |  |  |  |  |  |  |  |  |  | N/a | No | N/a | Yes | No |
| SHARP (quantitative) (17-20) |  |  |  |  |  |  |  |  |  |  | Yes | Yes | No | N/a | Yes |  |  |  |  |  |  |  |  |  |  |
| SHARP (qualitative) (21) | Yes | Yes | Yes | Yes | Yes |  |  |  |  |  |  |  |  |  |  |  |  |  |  |  |  |  |  |  |  |
| UK Millennium Cohort (22) |  |  |  |  |  |  |  |  |  |  | Yes | Yes | Yes | Yes | No |  |  |  |  |  |  |  |  |  |  |
| **FINANCIAL INCENTIVES** | | | | | | | | | | | | | | | | | | | | | | | | | |
| Section 8 (mixed) (23, 24) |  |  |  |  |  |  |  |  |  |  |  |  |  |  |  |  |  |  |  |  | Yes | Yes | Yes | Yes | N/a |
| Section 8 (RCT) (25) |  |  |  |  |  | N/a | Yes | Yes | No | N/a |  |  |  |  |  |  |  |  |  |  |  |  |  |  |  |
| Section 8 (panel study) (26) |  |  |  |  |  |  |  |  |  |  | No | Yes | N/a | Yes | No |  |  |  |  |  |  |  |  |  |  |
| Section 8 (cross-sectional) (27) |  |  |  |  |  |  |  |  |  |  |  |  |  |  |  | Yes | Yes | Yes | N/a | Yes |  |  |  |  |  |
| Norwegian Housing Allowance (28) |  |  |  |  |  |  |  |  |  |  |  |  |  |  |  | N/a | N/a | Yes | Yes | N/a |  |  |  |  |  |
| Spanish charity (29-31) |  |  |  |  |  |  |  |  |  |  | N/a | Yes | No | Yes | N/a |  |  |  |  |  |  |  |  |  |  |

Additional file 5 – Mixed Methods Appraisal Tool (MMAT) assessments

Appraisal Tool (MMAT)

RCT = randomised controlled trial; SHARP = Scottish Housing Health and Regeneration Project

### MMAT Criteria

**“**1. QUALITATIVE:

1.1. Is the qualitative approach appropriate to answer the research question?;

1.2. Are the qualitative data collection methods adequate to address the research question?;

1.3. Are the findings adequately derived from the data?;

1.4. Is the interpretation of results sufficiently substantiated by data?;

1.5. Is there coherence between qualitative data sources, collection, analysis and interpretation?

2. QUANTITATIVE (RCT):

2.1. Is randomization appropriately performed?;

2.2. Are the groups comparable at baseline?;

2.3. Are there complete outcome data?;

2.4. Are outcome assessors blinded to the intervention provided?;

2.5 Did the participants adhere to the assigned intervention?

3. QUANTITATIVE (NON-RANDOMISED):

3.1. Are the participants representative of the target population?;

3.2. Are measurements appropriate regarding both the outcome and intervention (or exposure)?;

3.3. Are there complete outcome data?;

3.4. Are the confounders accounted for in the design and analysis?;

3.5. During the study period, is the intervention administered (or exposure occurred) as intended?

4. QUANTITATIVE (DESCRIPTIVE):

4.1. Is the sampling strategy relevant to address the research question?;

4.2. Is the sample representative of the target population?;

4.3. Are the measurements appropriate?;

4.4. Is the risk of nonresponse bias low?;

4.5. Is the statistical analysis appropriate to answer the research question?

5. MIXED METHODS:

5.1. Is there an adequate rationale for using a mixed methods design to address the research question?;

5.2. Are the different components of the study effectively integrated to answer the research question?;

5.3. Are the outputs of the integration of qualitative and quantitative components adequately interpreted?;

5.4. Are divergences and inconsistencies between quantitative and qualitative results adequately addressed?;

5.5. Do the different components of the study adhere to the quality criteria of each tradition of the methods involved?
